# Novel coronavirus 2019-nCoV: early estimation of epidemiological parameters and epidemic predictions

**DOI:** 10.1101/2020.01.23.20018549

**Authors:** Jonathan M. Read, Jessica R.E. Bridgen, Derek A.T. Cummings, Antonia Ho, Chris P. Jewell

## Abstract

Since first identified, the epidemic scale of the recently emerged novel coronavirus (2019-nCoV) in Wuhan, China, has increased rapidly, with cases arising across China and other countries and regions. using a transmission model, we estimate a basic reproductive number of 3.11 (95%CI, 2.39–4.13); 58–76% of transmissions must be prevented to stop increasing; Wuhan case ascertainment of 5.0% (3.6–7.4); 21022 (11090–33490) total infections in Wuhan 1 to 22 January.

**Changes to previous version:**

- case data updated to include 22 Jan 2020; we did not use cases reported after this period as cases were reported at the province level hereafter, and large-scale control interventions were initiated on 23 Jan 2020;
- improved likelihood function, better accounting for first 41 confirmed cases, and now using all infections (rather than just cases detected) in Wuhan for prediction of infection in international travellers;
- improved characterization of uncertainty in parameters, and calculation of epidemic trajectory confidence intervals using a more statistically rigorous method;
- extended range of latent period in sensitivity analysis to reflect reports of up to 6 day incubation period in household clusters;
- removed travel restriction analysis, as different modelling approaches (e.g. stochastic transmission, rather than deterministic transmission) are more appropriate to such analyses.

## Background and current epidemic situation

On 29 December 2019, Chinese authorities identified a cluster of similar pneumonia cases of unknown aetiology in Wuhan City, Hubei Province, China (Tan *et al.*, 2020). A novel strain of coronavirus (2019-nCoV) was subsequently isolated from a patient on 7 January 2020 (Tan *et al*., 2020). Most cases from the initial cluster had epidemiological links with a live animal market (Huanan South China Seafood Market), suggesting a possible zoonotic origin (World Health Organization, 2020). However, the definitive source of the virus is unknown. Infections in at least one family cluster (Chan *et al*., 2020) and in healthcare workers confirm the occurrence of human-to-human transmission, though the extent of this mode of transmission is unclear. On 21 January 2020, the WHO suggested there was possible sustained human-to-human transmission (*World Health Organization Western Pacific on Twitter*, 2020).

As of 1000 GMT 25 Jan 2020, 1,372 cases had been confirmed (Munroe, 2020), of which more than 700 were from Hubei Province (BBC News, 2020). Cases have also been reported in other Chinese provinces, including large cities such as Beijing, Shanghai and Shenzhen, as well as other countries, including Thailand (n=5), Japan (n=2), South Korea (n=2), Taiwan (n=3), Malaysia (n=1), Singapore (n=3), Nepal (n=1), Vietnam (n=2), United States (n=2), Australia (n=1), and France (n=3) (European Centre for Disease Prevention and Control, 2020). In a case series of the first 41 hospitalised cases in Wuhan, presenting symptoms included fever, cough, myalgia and fatigue (Huang *et al*., 2020). Over half had shortness of breath, and all had evidence of pneumonia (Huang *et al*., 2020). To date, 41 deaths have been reported, all but two were from the Hubei province (European Centre for Disease Prevention and Control, 2020).

Current clinical and epidemiological data are insufficient to understand the full extent of the transmission potential of the epidemic. Estimates in this manuscript are highly uncertain due to uncertainty about the timing and natural history of cases, the impact of changes in reporting over the course of the outbreak and the potential impact of responses to cases among other sources. The outbreak comes at a time when there is a substantial increase in travel volume within as well as in and out of China around the Lunar New Year on 25 January 2019. Over 3 billion passenger journeys were predicted for the period between 10 January and 18 February (CGTN, 2020). In an effort to contain the outbreak, travel restrictions were imposed on Wuhan from 23 January, and have since expanded to 12 other cities, and large social gatherings cancelled (New York Times, 2020).

### Domestic and international connectivity of Wuhan

Wuhan is a city of more than 11 million residents and is connected to other cities in China via high-speed railway and frequent commercial airline flights. There were 670,417 airline passenger bookings departing Wuhan made during January 2017, the top destinations being Shanghai (53,214 bookings), Beijing (51,066 bookings) and Kunming (40,120 bookings) (OAG, 2020); **Figure 1**. While the majority of air travel departing Wuhan is domestic (87.2% of bookings, Jan 2017), Wuhan is connected internationally through both direct and indirect flights (Bogoch *et al*., 2020).

**Figure 1.**
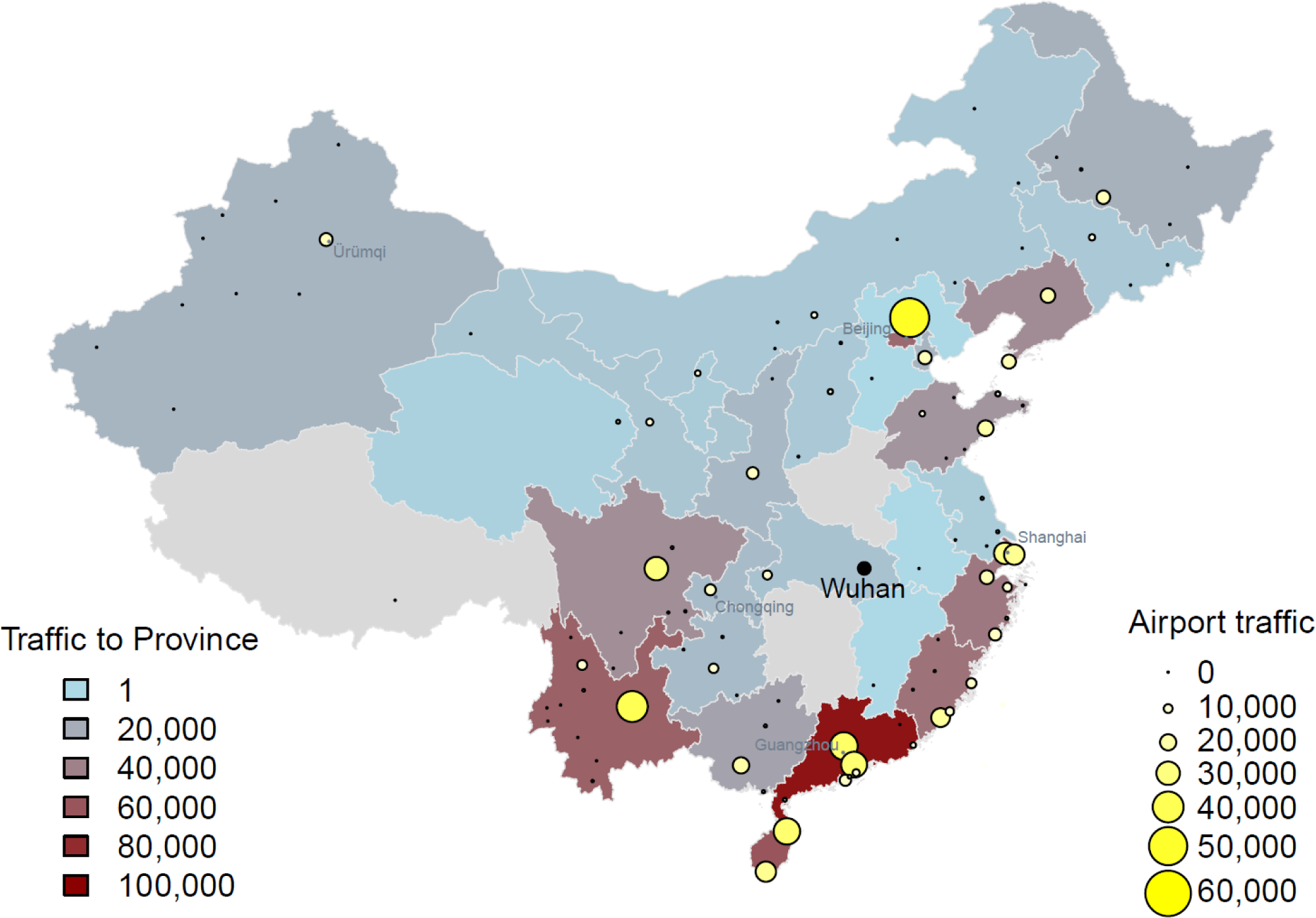
Connectivity of Wuhan to other cities and provinces in mainland China, based on total commercial airline traffic from Wuhan in January 2017. Traffic is based on the number of departing bookings.

### Transmission model

We fitted a deterministic SEIR metapopulation transmission model of infection within and between major Chinese cities to the daily number of confirmed cases of 2019-nCoV in Chinese cities and cases reported in other countries/regions, using an assumption of Poisson-distributed daily time increments (see Methods Supplement). We modelled the period from 1 January 2020 when local authorities closed the wet market implicated as the zoonotic source of human infection (World Health Organization, 2020) up to and including 22nd January 2020. We only considered human-to-human transmission in our model, and made the assumption that following the closure of the market, no further zoonotic infection contributed to the epidemic dynamics. Coupling between cities followed daily-adjusted rates of travel estimated from monthly-aggregated full itinerary passenger booking data for January 2017, accessed from OAG Traffic Analyser database (OAG, 2020), assuming that travellers are drawn randomly from the origin population.

We estimated the transmission rate and the recovery rate (the inverse of the infectious period). We assumed that the latent period was 4 days, based on an estimate of the incubation period of SARS, a related coronavirus (Lessler *et al*., 2009). This is similar to the estimate of 4.4 days from initial characterisation of 2019-nCoV cases. We make the assumption that the latent period approximates to the incubation period. We also estimated the ascertainment rate within Wuhan, and the initial number of human infections present in Wuhan when the market was closed. Confirmed cases in Chinese cities and other countries/regions reported as of 22 January 2020 were used for fitting; from 23 January cases were not reported for Wuhan and other locations within Hubei but only at the aggregate province level. Fitting was achieved by treating the ODE system as representing the mean number of new cases per day in our study period, and assuming that the observed number of new cases were (approximately) Poisson distributed around this mean. Given the model and data, parameter inference was achieved by maximum likelihood estimation using Nelder-Mead optimisation as implemented in the *optim()* function in the R statistical language (R Core Team 2020); see https://github.com/chrism0dwk/wuhan/tree/v0.2 for R code, case data, and prepared datafiles.

Uncertainty in the parameter estimates was explored using parametric bootstrap according to the following procedure. Firstly, 10000 Monte Carlo simulations from the model (ODE and Poisson noise) were generated using the MLE estimates of the parameters. Each simulated dataset was then re-fitted to the model to construct a joint sampling distribution of the parameters, and 95% confidence estimated as the lower 2.5% and upper 97.5% quantiles. The ODE system (without Poisson noise) was run over this sampling distribution to generate 95% confidence intervals around the predicted mean epidemic trajectory.

### Epidemiological parameter estimates

We estimated the transmission rate within Wuhan, *β*, to be 1.94 d^-1^ (95%CI, 1.25–6.71), while we found the infectious period to be 1.61 days (95%CI, 0.35–3.23). We calculated the basic reproductive number, *R*_0_, of the infection to be 3.11 (95%CI, 2.39–4.13), comparable to the range for SARS estimated from outbreaks during the 2003 epidemic (Lipsitch *et al*., 2003; World Health Organization, 2003). We highlight that this number is highly uncertain and that a large range of parameters are consistent with the data given the assumptions of our model. This estimate reflects both the dynamics of transmission and, potentially, the dynamics of case reporting, with increases as reporting over time potentially increasing our estimate. The estimate we give here represents an update from the report of 23 January in which we estimated a higher R, and highlights the sensitivity of our findings to additional data at this early period. The current estimate of *R*_0_is significantly greater than 1, the epidemic threshold, providing evidence that sustained human-to-human transmission is occurring in China. Further, it suggests that a concerted effort will be required to control the outbreak, requiring between 38% and 80% of transmission to be averted to control the epidemic.

We estimated that the average ascertainment rate in Wuhan between 1 and 22 January was 5.0% (95%CI, 3.6–7.4), reflecting the difficulty in identifying cases of a novel pathogen. Given the generally good accessibility to healthcare in China, this suggests that the majority of infections may be of mild illness and insufficiently serious for individuals to seek treatment. However, it is worth noting that a number of identified cases have died (Centre for Disease Control and Prevention 2020) and that the true case fatality rate has yet to be estimated accurately. Also, asymptomatic infection has been reported for 2019-nCoV (Chan *et al*., 2020). We also estimated the size of the epidemic in Wuhan at the time of the market closure (1 January) to be 15 individuals (95%CI, 5–37). Our estimates of epidemiological parameters are sensitive to our assumption regarding the length of the latent period; see **Figure 2**. Early epidemiological investigations suggest a duration between 3 to 6 days (Chan *et al*., 2020), Should the latent period be longer than the 4 days we assume, our *R*_0_ estimates would be higher and the estimated ascertainment rate slightly lower; see **Figure 2**. If cases are being reported with increasing efficiency and the timing of cases is inconsistent with the timing assumed here (i.e. throughout the outbreak, the length of time between infection and reporting in surveillance data is declining), this may tend to decrease our estimate of the reproductive number.

**Figure 2.**
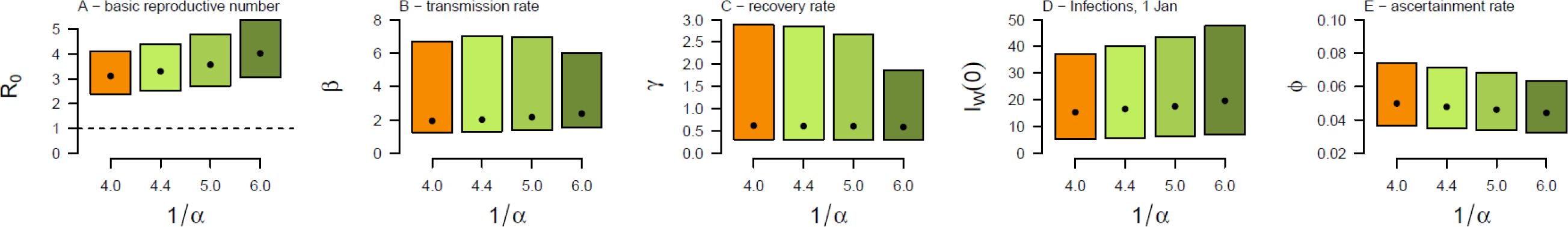
Sensitivity of parameter estimates to assumed latent period (1/*α*) value. Boxes represent the 2.5% and 97.5% quantiles and black dots the 50% quantile.

### Epidemic size estimates

Using our parameterised transmission model, we simulated the impact of an ongoing outbreak in Wuhan to seed infections and outbreaks in other cities of China, and to generate infection in travellers to other countries/regions, through airline travel originating in China. We stress that these projections make strong assumptions: that no control interventions are instigated; that the key epidemiological variables driving epidemic dynamics remain constant; that travel behaviour within China and to other countries/regions continues as per our mobility estimates; finally, we only consider travel by air and do not include land transportation, particularly via the rail network within China.

We estimate that on 22 January, in Wuhan there were currently 14464 infected individuals (prediction interval, 6510–25095), and a total 21022 infections (prediction interval, 11090– 33490) since the start of the year. We also estimate t 24 currently infected individuals (prediction interval, 19–30) in other locations of China on this date. For comparative purposes, we estimate the total number of infections in Wuhan from 1January to 18 January inclusive to have been 6733 (prediction interval, 3500–10914). This estimate of the total infections is comparable to other published estimates based on travel data and reported cases identified outside of China (estimated between 1,700 and 7,800)(Imai *et al*., 2020a), and highlights our estimated low ascertainment rate, the rapid growth of the epidemic, and uncertainty in model predictions.

Should the transmission continue at the same rate in Wuhan, with no control or change in the behaviour of individuals (such as spontaneous social distancing) our model predicts that on 29 January the epidemic in Wuhan will be substantially larger, with 594 cases expected to be detected on that day in Wuhan (prediction interval, 446–788) and 105077 currently infected (prediction interval, 46635–185412); see **Figure 3** and **Table 1**. If transmission has reduced, either through control or spontaneous public response to the epidemic, this will be a gross overestimate, though it may be useful to help gauge the effectiveness of interventions.

**Table 1.**
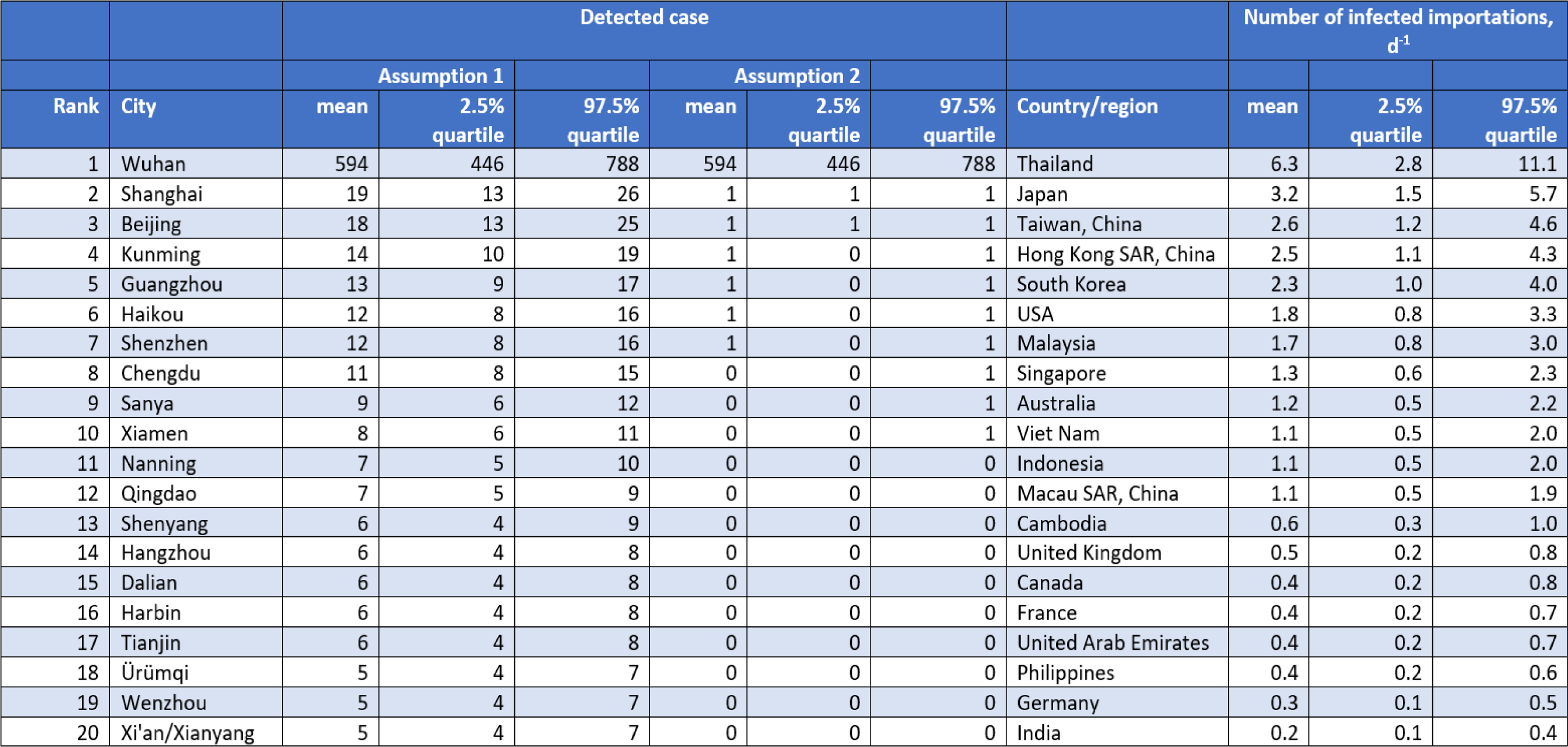
Predicted epidemic sizes (number of detected cases) in selected Chinese cities and predicted imports to other countries/regions on 29 January 2020, assuming no change in transmissibility or ascertainment rate. Assumption 1: ascertainment rate in all cities excluding Wuhan is 100%. Assumption 2: ascertainment rate in all Chinese cities is 5.0%.

**Figure 3.**
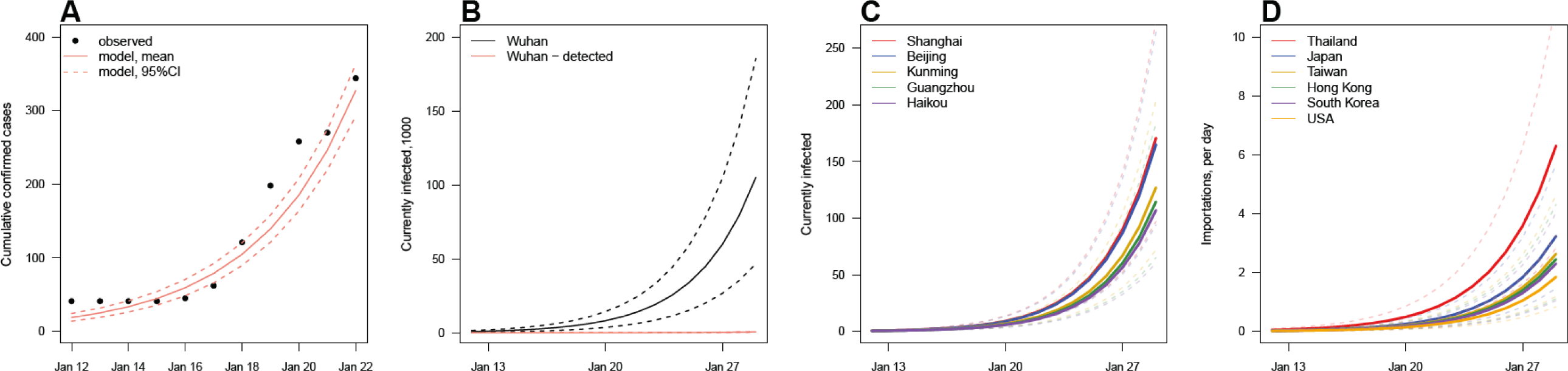
(**A**) Comparison of observed cases and predicted detected cases in Wuhan for the period 12 to 22 January. Epidemic predictions for (**B**) Wuhan, (**C**) selected Chinese cities and (**D**) selected countries/regions, up to 29 January. Estimated detected cases are also plotted for Wuhan. 95% confidence intervals around mean epidemic trajectory shown as dotted lines.

We also estimated epidemic projections for other Chinese cities and countries/regions. The epidemic sizes projections are subject to model and parameter uncertainty, though the relative magnitude of predictions in other cities and countries/regions may be informative for planning and preparedness purposes.The model predicts infected travellers to other Chinese cities will initiate outbreaks in those cities, the largest on 29 January being in Shanghai, Beijing, Guangzhou, Shenzhen, Chengdu and Kunming; see **Figure 3** and **Table 1**. Our model predicts the total number of infected individuals in locations elsewhere in China to be 237 (prediction interval, 167–324) on 29 January. Finally, the model predicts an elevated risk of importations into other countries/regions, most notably to Thailand, Japan, South Korea, Taiwan, Hong Kong SAR, USA, Singapore, Malaysia, Australia and Viet Nam; see **Figure 3** and **Table 1**. Again, these predictions assume no change in the transmission of the virus within China through control or other responses to the epidemic, and likely underestimate the potential importation rate to regions with ground transportation from China, in particular Hong Kong.

### Comparison of estimates to other reports

Our estimates of *R*_0_ are broadly consistent with early estimates from other groups: 2.0–3.3 (Majumder and Mandl, 2020); 2.6 (uncertainty range 1.5-3.5)(Imai *et al*., 2020b); 2.92 (95% CI 2.28, 3.67)(Liu *et al*., 2020); 2.2 (90% interval: 1.4-3.8) (Riou and Althaus, 2020). Sources of discrepancies may be due to model differences and differences in the contribution of specific types of data to our estimates. We believe that our estimates are slightly elevated compared to others due to the inclusion of cases from other locations within China other than Wuhan. However, it is important to note that our point estimate is consistent with all others uncertainty intervals, all indicating sustained growth of cases.

### Comparison of transmissibility with SARS and MERS

Our estimates of the basic reproductive number for this novel coronavirus are comparable to most estimates reported for SARS and MERS-CoV, but similar to some estimates from subsets of data in the early period of SARS. For the SARS coronavirus, estimates of the mean reproductive number ranged from 1.1 to 4.2 with most estimates between 2 and 3 (Bauch *et al*., 2005). These estimates represent a range of methods and settings. Some estimates come from data that mixes time periods before and after control. Estimates of *R*_0_ also varied based on assumed serial intervals (e.g. (Lipsitch *et al*., 2003) estimated *R*_0_ ranging from 2.2 to 3.6 for serial intervals of 8 to 12 days (9). Another study (Bauch *et al*., 2005) reviewed sources of variation in basic reproductive numbers of SARS and noted that those locations in which outbreaks occurred, *R*_0_ was approximately 3. Estimates from MERS-CoV were uniformly lower, with estimates from Saudi Arabia having a mean of less than 1 (∼0.5) but exhibited large temporal variability with increases in some periods of time particularly in healthcare settings (Cauchemez *et al.*, 2016).

A comparison of the efficiency of transmission in this outbreak and in SARS outbreaks can be seen as well in simple comparisons of doubling times in each outbreak. In SARS, doubling times varied from 4.6 days to 14.2 days depending on setting (doubling time, Td=6.0 (1358 over 63 days, Singapore), Td=4.6 (425 over 41 days, Hong Kong), Td=14.2 (7919 over 185 days, overall)) (Lipsitch *et al*., 2003). Using confirmed case information (41 reported January 14; 291 reported 24:00 on January 20; 1975 reported 24:00 on January 25)(*NHCPRC daily reports*, 2020) we find doubling times of 2.1, 1.8, and 2.0 days. If the outbreak has been ongoing for a longer period of time, this would increase the estimated doubling time. These doubling time estimates, similar to our estimates of *R*_0_, are susceptible to bias due to the dynamics of case reporting, with bunching of identified cases (due to temporally clustered recognition of cases) tending to bias our estimate towards lower doubling times. We note our estimates of the doubling time in this outbreak are short compared to estimates from the SARS outbreak in Hong Kong (Lipsitch *et al.*, 2003).

## Limitations

Our model necessarily makes a number of assumptions. Our estimates of the basic reproductive number of this novel coronavirus are tied to the specific time period and data analysed here, and this measure may change substantially over the course of this outbreak and as additional data arrives. Additionally, the spatial component of our model is dependent upon only airline travel; the model does not include rail and road transportation, so we may underestimate local connectivity and the connectivity of Wuhan to other locations. We also do not attempt to account for any implementation of control, nor and dynamic changes of factors than may influence transmission (such as spontaneous social distancing), nor changes in surveillance and reporting effort. Our choice of modelling approach may also lead to unreliability in the precision of our estimated model parameters (King *et al*. 2015), However, our approach used ‘raw’ counts of cases to fit the model, not cumulative case information, and the point estimates would not be biased (King *et al*. 2015).

Earlier novel coronavirus (SARS and MERS-CoV) outbreaks found evidence for substantial heterogeneity in reproductive numbers between individuals (Chowell *et al*., 2004; Bauch *et al.*, 2005; Cauchemez *et al.*, 2016). In our analysis, we assume that there is little heterogeneity in reproductive numbers and this assumption may change our estimated reproductive number. Additionally, *R*_0_ estimates tend to be reduced as case information accumulates, though control measures may also be introduced during these periods. As is true for any modelling analysis of surveillance data, our estimate of *R*_0_ may also reflect the dynamics of surveillance effort and reporting rather than just the dynamics of the epidemic.

A key uncertainty of this outbreak is when it started. We have chosen to model transmission from 1 January onwards. Surveillance in China and elsewhere only started once the outbreak was identified in Wuhan. Had the outbreak started much earlier, and both within-China and international infectious exports occurred before January and in early January (while surveillance was ramping up), our estimates of the reproductive number would mostly decrease.

A threat to the accuracy of these projections is if a substantial proportion of infection has been due to multiple exposures to animals that has been curtailed in some way. These data may also represent a period of high transmission (due to favourable seasonal conditions, stochastic variation or selection bias in detecting large clusters of transmission) that will not be sustained over long periods of time.

## Summary

We are still in the early days of this outbreak and there is much uncertainty in both the scale of the outbreak, as well as key epidemiological information regarding transmission. However, the rapidity of the growth of cases since the recognition of the outbreak is much greater than that observed in outbreaks of either SARS or MERS-CoV. This is consistent with our broadly higher estimates of the reproductive number for this outbreak compared to these other emergent coronaviruses, suggesting that containment or control of this pathogen may be substantially more difficult.

## Data Availability

The collated case and population data used for modelling is provided at a linked repository. Domestic and international airline passenger data are available via subscription from OAG (www.oag.com).

https://github.com/chrism0dwk/wuhan/tree/v0.2

## Notes

### Competing Interest Statement

The authors have declared no competing interest.

### Funding Statement

JMR and CPJ acknowledge support from the Medical Research Council (MR/5004793/1). JMR acknowledges support from the Engineering and Physical Sciences Research Council (EP/N014499/1). JREB acknowledges support from Faculty of Health and Medicine, Lancaster University in the form of a PhD Scholarship. CPJ acknowledges support from Wellcome.

